# Neonatal Adiposity is Largely Driven by Maternal Hyperglycemia *in utero*: Analysis of Routine Clinical Data from a Diabetes Clinic in India

**DOI:** 10.1101/2024.04.27.24306016

**Authors:** Sayali S. Deshpande-Joshi, Sonali S. Wagle-Patki, Madhura K. Deshmukh, Himangi G. Lubree, Hemant S. Damle, Suhas R. Otiv, Sanat B. Phatak, Rucha H. Wagh, K. Meenakumari, Smita N. Dhadge, Rajashree P. Kamat, Sayali G. Wadke, Deepa A. Raut, Dattatray S. Bhat, Souvik Bandyopadhyay, Chittaranjan S. Yajnik

## Abstract

**Aim:** To examine the effect of maternal size and degree of hyperglycemia on neonatal adiposity in the relatively thin Indian population.

**Study design:** Analysis of routine clinical data collected in one diabetes clinic.

**Methods:** We examined the association of maternal size (BMI) and degree of hyperglycemia (type of diabetes, type 1 being the thinnest and most hyperglycemic, type 2 and Gestational diabetes being overweight and obese but less hyperglycemic) with neonatal adiposity measurements (weight, ponderal index, abdominal circumference, and skinfold thickness) using multiple linear regression.

**Results:** We included data on 772 pregnancies with diabetes (61 with type 1, 79 with type 2, and 632 with gestational) and 349 with normal glucose tolerance (NGT). Mothers with type 1 diabetes had the lowest BMI and highest HbA1c, however, their neonates were the most obese, centrally obese, and adipose. Compared to neonates of NGT mothers, those of mothers with type 1 diabetes were 370 g heavier, those of mothers with type 2 diabetes 265 g, and those of mothers with GDM by 200 g. Prediction models adjusted for gestational age at birth, neonatal sex, maternal age, parity, and year of birth confirmed that neonates of mothers with type 1 diabetes were the most adipose (ponderal index, abdominal circumference and skinfolds), followed serially by those of mothers with type 2 diabetes, GDM, and NGT. Compared to maternal type of diabetes, pre-pregnancy BMI and gestational weight gain (GWG) made a much smaller contribution to neonatal adiposity.

**Conclusion:** Our findings provide a clear answer that maternal hyperglycemia rather than BMI is the primary driver of neonatal adiposity. Adequate control of maternal hyperglycemia will help control neonatal adiposity.

**RESEARCH IN CONTEXT:** *What is already known about this subject?:*

- Maternal obesity and diabetes increase neonatal adiposity which contributes to bad pregnancy outcomes
- Given the strong association between maternal obesity and diabetes in western populations, the dominant determinant of neonatal adiposity is difficult to assess

*What is the key question?:*

Is maternal BMI or hyperglycemia the primary determinant of neonatal adiposity?

*What are the new findings?:*

- Indian mothers with type 1 diabetes were the thinnest and the most hyperglycemic compared to mothers with type 2 diabetes and GDM who were more obese and less hyperglycemic, providing a contrasting exposure
- Neonates of mothers with type 1 diabetes were the most obese-adipose, those of mothers with type 2 diabetes and GDM were progressively less so.

*How might this impact on clinical practice in the foreseeable future?:*

- Our results clearly show that maternal hyperglycemia is the major driver of neonatal adiposity. Strict control of maternal hyperglycemia will help reduce neonatal adiposity and associated adverse outcomes

## INTRODUCTION

In 2021, an estimated 16.7% of live births worldwide (> 20 million) were exposed to some form of glucose intolerance in pregnancy. Of these, 10-20% had pre-gestational diabetes (type 1, type 2, and other) while the majority (80%) had gestational diabetes [1]. Maternal diabetes during pregnancy is associated with adverse short- and long-term outcomes for the offspring [2]. The main short-term effect is excessive fetal growth and its sequelae in the peripartum period (difficult labour, neonatal injuries, need for interventional delivery, post-natal complications etc.). Pedersen proposed from his observations in pregnancies in (lean) women with type 1 diabetes that the transfer of excess maternal glucose to the fetus stimulates fetal islets, resulting in hyperinsulinemia and macrosomia [3]. Freinkel, studying a population of more obese women with diabetes proposed that an excess transfer of a “mixture” of maternal nutrients (glucose, lipids, and amino acids) in pregnancies with diabetes promotes fetal macrosomia and long-term risk of obesity and diabetes (fuel mediated teratogenesis) [4]. Thus, maternal diabetes has been proposed to contribute to the cascading epidemic of obesity-adiposity and diabetes [5–7]. Maternal obesity, independent of glycaemic status is also known to contribute to neonatal adiposity [8–11]. There is a broad agreement that maternal diabetes and obesity concomitantly increase the risk for neonatal adiposity, however, there has been no attempt to investigate the relative influence of these two exposures. In populations from high-income countries, maternal diabetes and obesity co-exist, making it difficult to distinguish individual contributions of these two related yet distinct exposures on offspring adiposity. Clarity about the relative effects of maternal diabetes and obesity may come from studies in populations where these two characteristics are dissociated.

At the Diabetes Unit, King Edward Memorial Hospital Research Centre (KEMHRC), Pune, we have collected data on a substantial number of pregnancies in women with type 1 diabetes, type 2 diabetes and GDM over the last three decades. In our clinic, mothers with type 1 diabetes are the thinnest and most hyperglycaemic, while those with type 2 diabetes and GDM are heavier and less hyperglycaemic, providing a contrasting exposure. These unique combinations of phenotype provided an opportunity to answer the question: Is maternal BMI or glycemia the primary driver of neonatal adiposity?

## METHODS

### Study population

This is an analysis of contemporary clinical data from the Diabetes Unit, KEMHRC, Pune. We reviewed clinical records of 1223 pregnant women (and their neonates) who attended the diabetes clinic between 1986-2020 for treatment and management of diabetes (type 1 diabetes, type 2 diabetes, and GDM) during pregnancy. Women with pre-gestational diabetes were either registered with us for regular treatment from before pregnancy or were referred for management during pregnancy by practicing physicians. We also reviewed records of 463 normal glucose tolerant (NGT) women (75g OGTT) who participated in two observational studies in our department: 1) International Atomic Energy Agency (IAEA-B12) study (2004-06), which investigated vit. B12 metabolism during pregnancy [12], and 2) InDiaGDM (2014-16), a study of gestational diabetes. Inclusion criteria were: maternal age greater than 18 yrs, singleton pregnancy, gestational age at delivery between 32 and 42 weeks, and live births. Control data of NGT pregnancies was available for the two decades (2000-2010 and 2011-2020); therefore, we excluded data collected before the year 2000 in pregnancies with diabetes. Those with incomplete data and biologically impossible outliers were also excluded from the analysis (Supplementary Figure 1).

### Exposures

Maternal BMI after delivery and diabetes during pregnancy (none, type 1diabetes, type 2 diabetes and GDM) were the two main exposures. Maternal diabetes type was considered a surrogate for the degree of maternal hyperglycemia and her obesity/adiposity status: type 1 diabetes being the most hyperglycaemic but least obese-adipose (lowest BMI, waist circumference and skinfolds), type 2 diabetes intermediate and GDM the least hyperglycaemic but most obese-adipose. Paternal data was not available in this study.

Information obtained from the clinical records included: type of maternal diabetes, demographics, obstetric history, pregnancy complications, treatment, and delivery details (sex of the neonate, gestational age at delivery, mode of delivery, etc.). Maternal type 1 diabetes and type 2 diabetes were defined by the ADA clinical criteria [13]. GDM was diagnosed by a fasting 75 g OGTT (WHO 1999 criteria [Fasting Plasma Glucose (FPG) ≥ 6.1 mmol/L and/ 2-hr plasma glucose ≥ 7.7 mmol/L] till 2012 [14], and International Association of Diabetes and Pregnancy Study Groups (IADPSG) criteria thereafter (FPG ≥ 5.1 mmol/L or 2-hr glucose ≥ 8.5 mmol/L) [15].

Mothers with type 1 diabetes were treated with multiple daily doses of insulin, none had used insulin pump. Mothers with type 2 diabetes and GDM were treated with lifestyle advice, and oral anti-diabetic drugs (OADs, metformin or acarbose) and insulin if necessary. Maternal weight, height, waist circumference, biceps, triceps, subscapular and suprailiac skinfold thicknesses were measured by trained staff within 72 hours of delivery using a standard protocol [16] (Supplemental Table S1). Similar information was also available in NGT mothers.

### Outcomes

The outcomes included neonatal weight, generalized adiposity (ponderal index), central adiposity (abdominal circumference), and subcutaneous adiposity (sum of skinfold thickness). Trained research staff performed detailed anthropometry (weight, crown heel length, head circumference, abdominal circumference, triceps, and subscapular skinfold thicknesses) within 72 hours of birth using standardized protocols [16] (Supplemental Table S1). Neonatal adiposity measurements are expressed both as: 1) study specific SD scores calculated by the residual method on combined data from pregnancies with diabetes and without diabetes, adjusting for gestational age at birth and sex of the neonate. 2) by INTERGROWTH standards (2014) [17].

### Laboratory methods

Venous plasma glucose was measured using glucose oxidase-peroxidase method (Hitachi 902, Roche Diagnostics GmbH, Germany). Measurements were subject to external quality control (EQAS) and had CV <5%. HbA1c was measured chromatographically on Biorad D10 machine (Hercules, California).

### Statistical methods

We assessed the power of our study retrospectively for the outcomes of neonatal adiposity measures. Studying 772 pregnancies with diabetes and 349 with NGT would provide 90% power to detect a difference of 0.2 SD between the two groups at 5% significance.

Given the long duration of the studies, the criteria for diagnosis of GDM changed in our centre in 2013 from WHO (1985) to IADPSG (2010) criteria. Therefore, we created a categorical variable to represent these two time groups (births between 2000-2012 and 2013-2020)’ as a covariate to account for the variation contributed by different diagnostic criteria. We are aware that this variable may also reflect the improvement in different aspects of the management of pregnancies with diabetes. Over this long study duration, some data were missing; we report number of available observations for each variable.

Data are presented as median (25^th^ and 75^th^ percentile) or mean (± SD). Comparisons between groups were made by the Mann Whitney’s test or ANOVA (adjusting for gestational age at birth and sex in neonates). Differences in proportions were tested using the chi-square test. The association between maternal type of diabetes and neonatal adiposity was examined using multiple linear regression analysis, considering NGT mothers as the reference. These models were adjusted for maternal age at delivery and parity, and gestational age at birth, and sex of the neonate. Birth weight and other neonatal outcomes predicted from these models were plotted for the usual range of gestational age (32 - 42 weeks), using predictive modelling.

In addition to maternal BMI after delivery, we had data on maternal pre-pregnancy weight (n=390, self-reported) and thus GWG was calculated. We performed step-wise linear regression to assess the relative contribution of maternal factors to neonatal adiposity. Neonatal adiposity measures were dependent variables, each of the independent variable was included in the step-wise regression models to examine the R^2^ change; these included maternal pre-pregnancy BMI, gestational weight gain (GWG) and type of diabetes. These models were adjusted for maternal age, parity, gestational age at delivery and neonatal sex. We used SPSS version 21.0 (IBM corporation, Armonk, NY) and R-studio for data analysis. The analysis was performed during 2022-2023.

### Ethics

Use of the clinical data for research analysis was approved by KEMHRC Pune, Ethics Committee, (DIP 2128/08-12-2021). IAEA-B12 study (064/06-03-2006/15382/R0) and Indo-Danish study (BT/IN/Denmark/02/CSY/2014) also had KEMHRC Ethics Committee approvals. Indo-Danish study was registered with the Clinical Trials Registry-India (NCT03388723). All participants signed an informed consent before participation.

## RESULTS

Supplementary Figure 1 shows the flow chart of pregnancies that were considered for this analysis and the reasons for exclusion at each stage. There were no differences in age, anthropometry, and HbA1c measurements between included and excluded mothers (Supplemental Table S2), there were no differences in the included and excluded neonates for sex ratio and anthropometric measurements; however, the gestational age at birth was higher in included than those excluded (by three days, p<0.001). Thus, we included 772 pregnancies with diabetes (61 type 1 diabetes, 79 type 2 diabetes, and 632 GDM) and 349 NGT pregnancies on whom relevant data was available.

For the additional analysis, we considered maternal BMI after delivery, maternal pre-pregnancy weight (n=390, self-reported) and gestational weight gain (GWG) as independent variables. We investigated their contribution to neonatal adiposity. Mothers on whom this data was available were similar in age, weight, height, and BMI but had marginally higher HbA1c measurements compared to those on whom the data was not available (Supplemental Table S3). The neonates born to these two groups of mothers were similar in terms of anthropometric measurements.

### Mothers

Mothers with diabetes were older and had higher BMI, waist circumference, and skinfold thickness compared to NGT mothers (Table 1, Figure 1). Within the diabetes group, mothers with type 1 diabetes were the youngest, thinnest, and least adipose (central and subcutaneous adiposity). They also had the highest HbA1c percent, and longer duration of diabetes. All mothers with type 1 diabetes were treated with multiple daily injections of insulin. In mothers with type 2 diabetes, in addition to lifestyle adjustments, 50 % received insulin alone, 43% received insulin and OADs. Of the GDM mothers, in addition to lifestyle adjustment, 63% received insulin alone, 20% received insulin and OADs, 10% received OADs alone. Preterm and caesarean deliveries were more common in pregnancies with diabetes compared to NGT, especially in women with type 1 diabetes. None of the mothers in our study admitted to smoking.

**Figure 1.**
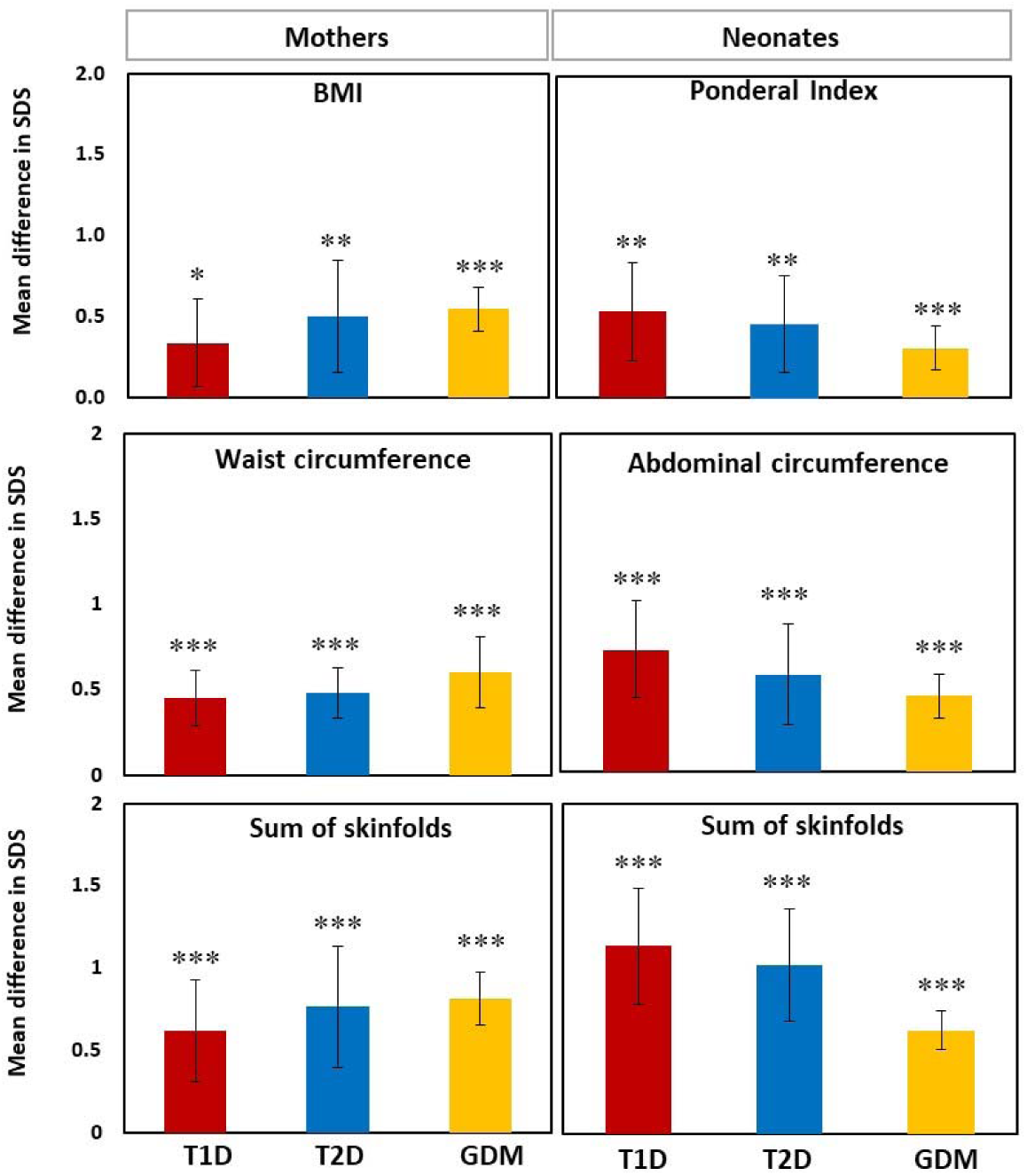
Adiposity in mothers and neonates. Figure shows adiposity indicators in mothers (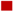 Type 1 diabetes 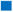 Type 2 diabetes 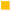 GDM) and their neonates in comparison with NGT. Height of the bars represents mean difference in SD score of each measurement from the NGT group (represented as zero line). * p<0.05, **p<0.01, ***p<0.001 using t-test.

**Table 1.**
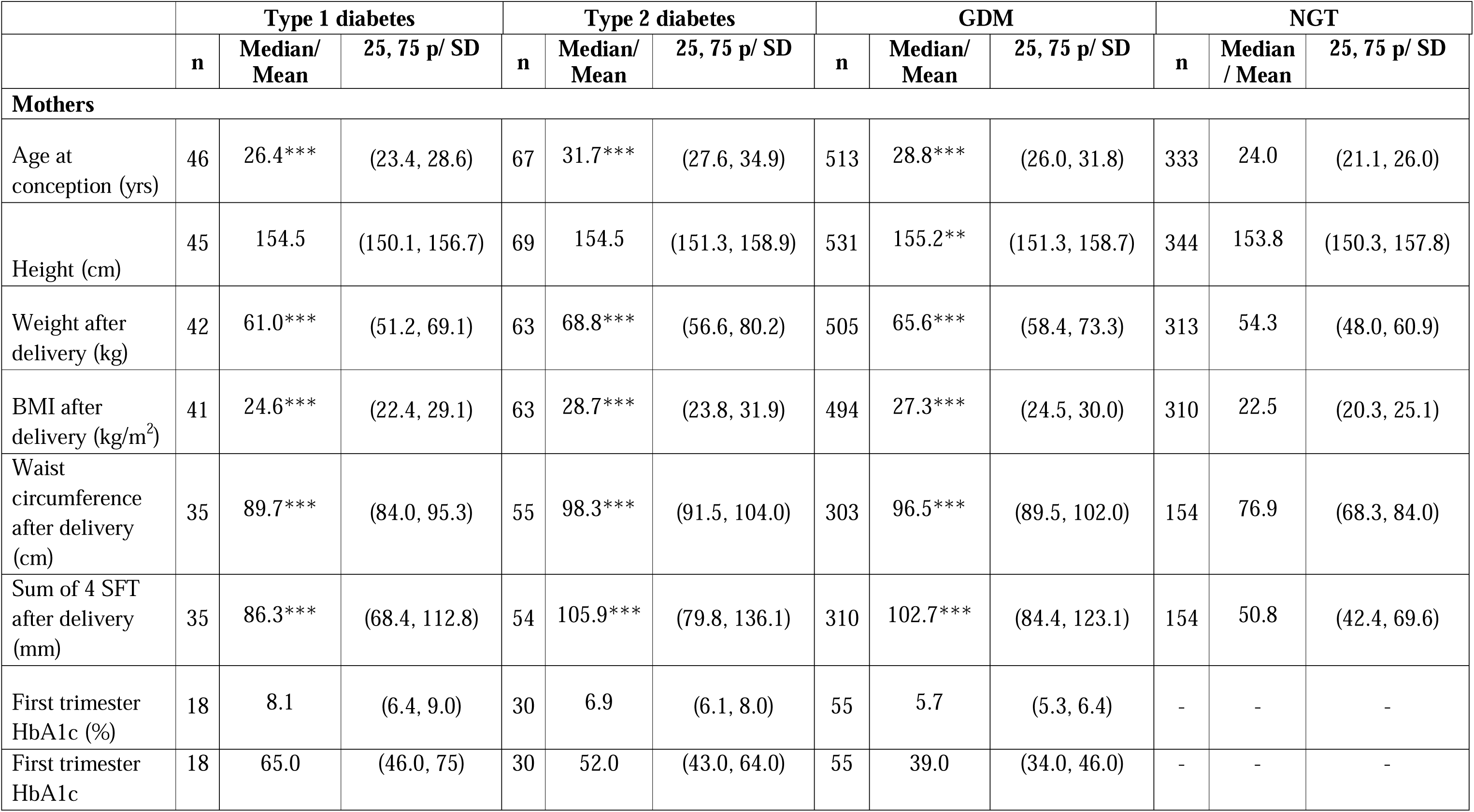
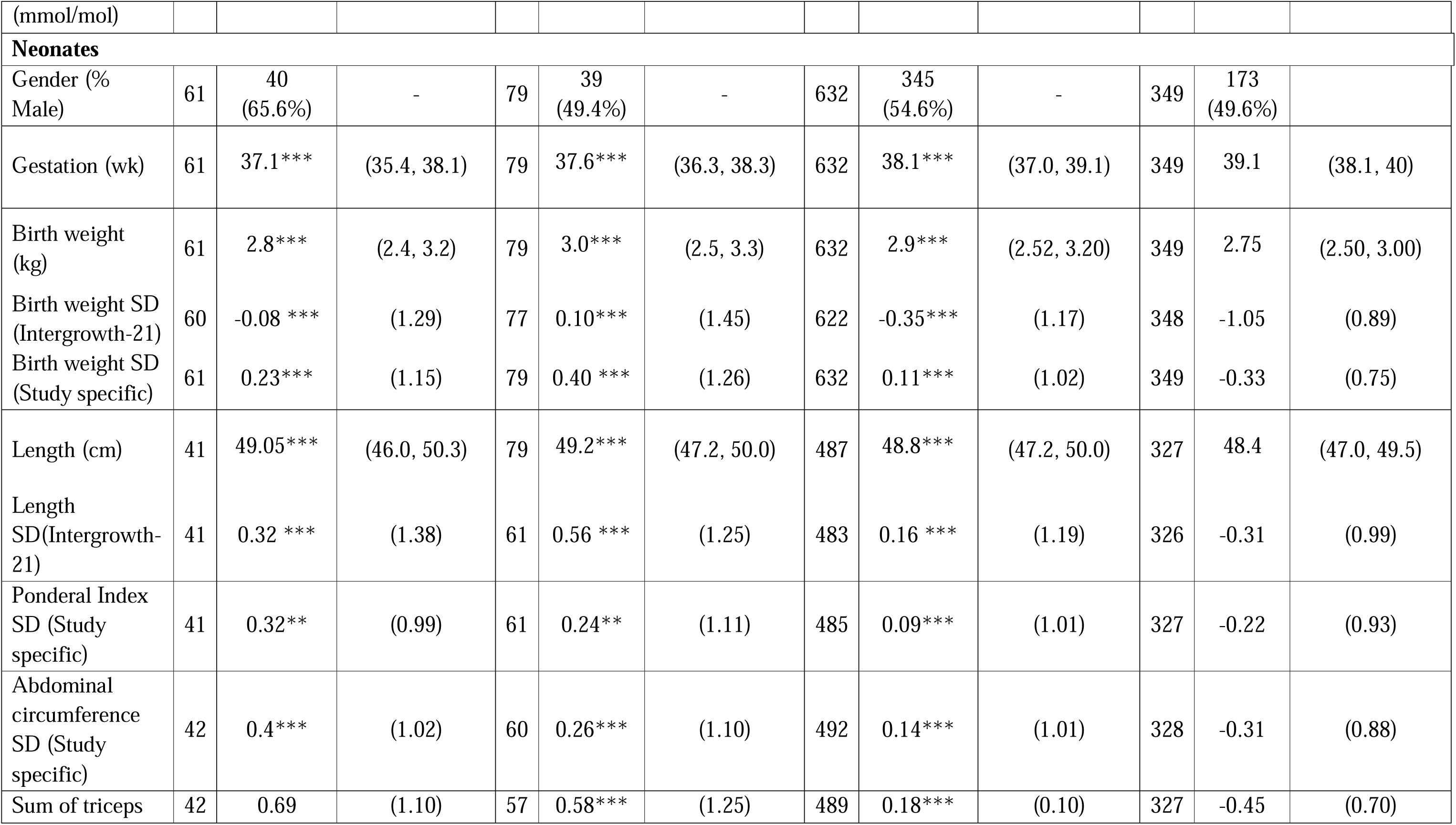
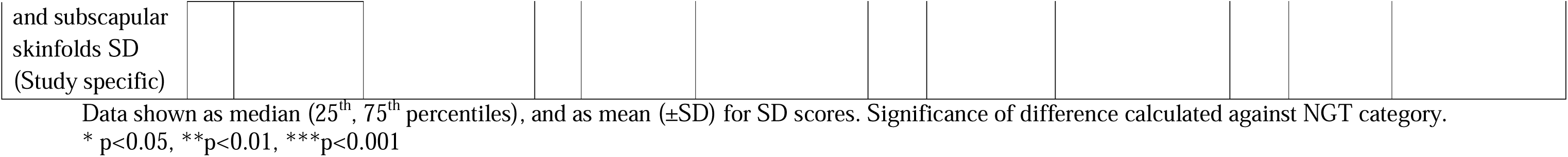
Maternal and neonatal characteristics.

|  | Type 1 diabetes |  |  | Type 2 diabetes |  |  | GDM |  |  | NGT |  |  |
| --- | --- | --- | --- | --- | --- | --- | --- | --- | --- | --- | --- | --- |
|  | n | Median/<br>Mean | 25, 75 p/ SD | n | Median/<br>Mean | 25, 75 p/ SD | n | Median/<br>Mean | 25, 75 p/ SD | n | Median<br>/ Mean | 25, 75 p/ SD |
| **Mothers** |  |  |  |  |  |  |  |  |  |  |  |  |
| Age at conception (yrs) | 46 | 26.4*** | (23.4, 28.6) | 67 | 31.7*** | (27.6, 34.9) | 513 | 28.8*** | (26.0, 31.8) | 333 | 24.0 | (21.1, 26.0) |
| Height (cm) | 45 | 154.5 | (150.1, 156.7) | 69 | 154.5 | (151.3, 158.9) | 531 | 155.2** | (151.3, 158.7) | 344 | 153.8 | (150.3, 157.8) |
| Weight after delivery (kg) | 42 | 61.0*** | (51.2, 69.1) | 63 | 68.8*** | (56.6, 80.2) | 505 | 65.6*** | (58.4, 73.3) | 313 | 54.3 | (48.0, 60.9) |
| BMI after delivery (kg/m<sup>2</sup>) | 41 | 24.6*** | (22.4, 29.1) | 63 | 28.7*** | (23.8, 31.9) | 494 | 27.3*** | (24.5, 30.0) | 310 | 22.5 | (20.3, 25.1) |
| Waist circumference after delivery (cm) | 35 | 89.7*** | (84.0, 95.3) | 55 | 98.3*** | (91.5, 104.0) | 303 | 96.5*** | (89.5, 102.0) | 154 | 76.9 | (68.3, 84.0) |
| Sum of 4 SFT after delivery (mm) | 35 | 86.3*** | (68.4, 112.8) | 54 | 105.9*** | (79.8, 136.1) | 310 | 102.7*** | (84.4, 123.1) | 154 | 50.8 | (42.4, 69.6) |
| First trimester HbA1c (%) | 18 | 8.1 | (6.4, 9.0) | 30 | 6.9 | (6.1, 8.0) | 55 | 5.7 | (5.3, 6.4) | - | - | - |
| First trimester HbA1c | 18 | 65.0 | (46.0, 75) | 30 | 52.0 | (43.0, 64.0) | 55 | 39.0 | (34.0, 46.0) | - | - | - |
| (mmol/mol) |  |  |  |  |  |  |  |  |  |  |  |  |
| **Neonates** |  |  |  |  |  |  |  |  |  |  |  |  |
| Gender (% Male) | 61 | 40<br>(65.6%) | - | 79 | 39<br>(49.4%) | - | 632 | 345<br>(54.6%) | - | 349 | 173<br>(49.6%) |  |
| Gestation (wk) | 61 | 37.1*** | (35.4, 38.1) | 79 | 37.6*** | (36.3, 38.3) | 632 | 38.1*** | (37.0, 39.1) | 349 | 39.1 | (38.1, 40) |
| Birth weight (kg) | 61 | 2.8*** | (2.4, 3.2) | 79 | 3.0*** | (2.5, 3.3) | 632 | 2.9*** | (2.52, 3.20) | 349 | 2.75 | (2.50, 3.00) |
| Birth weight SD (Intergrowth-21) | 60 | -0.08 *** | (1.29) | 77 | 0.10*** | (1.45) | 622 | -0.35*** | (1.17) | 348 | -1.05 | (0.89) |
| Birth weight SD (Study specific) | 61 | 0.23*** | (1.15) | 79 | 0.40 *** | (1.26) | 632 | 0.11*** | (1.02) | 349 | -0.33 | (0.75) |
| Length (cm) | 41 | 49.05*** | (46.0, 50.3) | 79 | 49.2*** | (47.2, 50.0) | 487 | 48.8*** | (47.2, 50.0) | 327 | 48.4 | (47.0, 49.5) |
| Length SD(Intergrowth-21) | 41 | 0.32 *** | (1.38) | 61 | 0.56 *** | (1.25) | 483 | 0.16 *** | (1.19) | 326 | -0.31 | (0.99) |
| Ponderal Index SD (Study specific) | 41 | 0.32** | (0.99) | 61 | 0.24** | (1.11) | 485 | 0.09*** | (1.01) | 327 | -0.22 | (0.93) |
| Abdominal circumference SD (Study specific) | 42 | 0.4*** | (1.02) | 60 | 0.26*** | (1.10) | 492 | 0.14*** | (1.01) | 328 | -0.31 | (0.88) |
| Sum of triceps | 42 | 0.69 | (1.10) | 57 | 0.58*** | (1.25) | 489 | 0.18*** | (0.10) | 327 | -0.45 | (0.70) |

### Neonates

As a group, neonates born in pregnancies with diabetes had higher birth weight, length, ponderal index, abdominal circumference, and skin fold thickness compared to those born in NGT pregnancies (Table 1, Figure 1). Their gestational age at delivery was lower. They also had a higher prevalence of large for gestational age (LGA; 10.4% vs. 0.3%) and a lower prevalence of small for gestational age (SGA; 22.1% vs. 41.4%). Offspring of mothers with type 1 diabetes had the highest ponderal index, abdominal circumference, and skin fold thickness, followed by offspring of mothers with type 2 diabetes and GDM (Figure 1).

### Maternal diabetes type, BMI, and neonatal adiposity

Figure 2 shows the simultaneous influence of the type of maternal diabetes and her BMI on neonatal adiposity measurements. Neonates of mothers with type 1 diabetes and high BMI had highest weight, abdominal circumference, and skinfold thickness.

**Figure 2:**
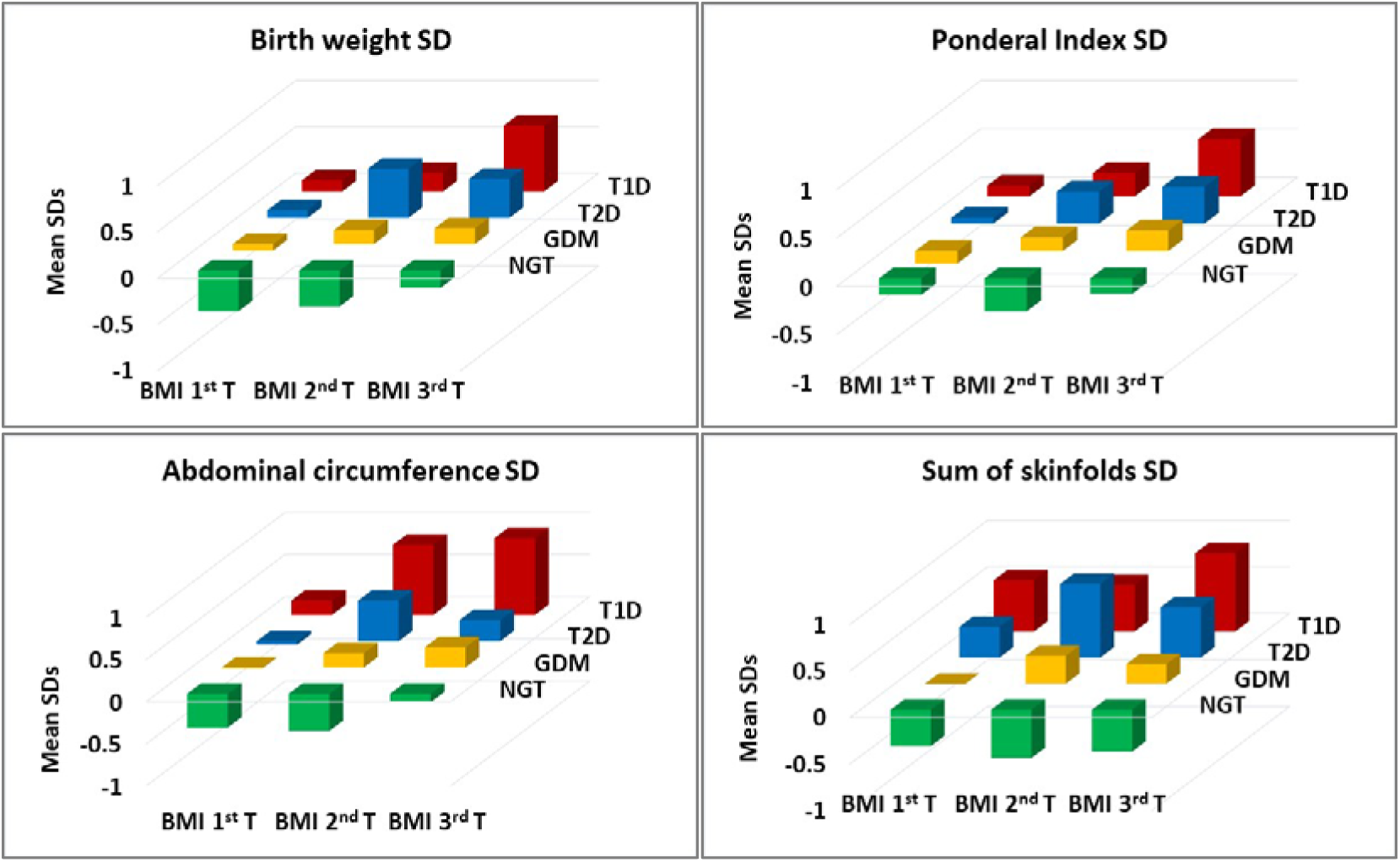
Mean SD scores of body size measurements in neonates by maternal glycemia (type 1 diabetes, type 2 diabetes, GDM, and NGT) and maternal BMI. Figure shows the distribution of mean SD scores of different body size measurements in neonates by maternal glycemia (type 1 diabetes, type 2 diabetes, GDM, and NGT) and maternal BMI (BMI T denotes tertiles (1^st^, 2^nd,^ and 3^rd^). Height of the bars represents mean SD scores.

### Quantifying the influence of maternal type of diabetes on neonatal adiposity

We performed multiple linear regression analysis to quantify the influence of type of maternal diabetes on neonatal size measurements, adjusting for maternal age and parity, neonatal gestational age at birth and sex, and calendar years (2000-2012 and 2013-2020). Neonates born to mothers with type 1 diabetes were 370 g heavier, had 0.15 g/cm^3^ higher ponderal index, 1.7 cm larger abdominal circumference, and 2.4 mm greater sum of skinfolds, compared to those born to mothers with NGT (Table 2). Maternal type 2 diabetes and GDM had a progressively lower influence as shown in the table. All the results were similar when only full-term (gestational age 37 to 42 weeks) babies were considered (n=886). (Supplemental Table S4).

**Table 2.** Multiple linear regression results to study association between maternal type of diabetes and neonatal adiposity measurements.

|  | Birth weight (g)<br>(n=890) |  | Ponderal Index<br>(g/cm<sup>3</sup>) (n=590) |  | Abdominal<br>circumference<br>(cm) (n=595) |  | Sum of skinfolds<br>(mm) (n=581) |  |
| --- | --- | --- | --- | --- | --- | --- | --- | --- |
|  | Estimate | SE | Estimate | SE | Estimate | SE | Estimate | SE |
| **Type of maternal diabetes** |  |  |  |  |  |  |  |  |
| Nil (NGT) | 0 |  | 0 |  | 0 |  | 0 |  |
| Type 1 | 370.5*** | 75.2 | 0.15*** | 0.05 | 1.73*** | 0.36 | 2.43*** | 0.37 |
| Type 2 | 265.4*** | 67.3 | 0.12** | 0.05 | 0.92*** | 0.32 | 1.82*** | 0.34 |
| GDM | 200.5*** | 36.8 | 0.10*** | 0.02 | 0.99*** | 0.17 | 1.33*** | 0.18 |
| **Maternal age at conception (yrs.)** | 2.0 | 3.7 | -0.00 | 0.00 | -0.00 | 0.02 | 0.03 | 0.02 |
| **Gestational age (wk)** | 131.4*** | 8.7 | 0.03*** | 0.01 | 0.39*** | 0.04 | 0.15*** | 0.00 |
| **Sex of neonate** |  |  |  |  |  |  |  |  |
| Male | 0 |  | 0 |  | 0 |  | 0 |  |
| Female | -65.2** | 29.3 | 0.03 | 0.02 | 0.02 | 0.14 | 0.68*** | 0.14 |
| **Parity** | 77.6*** | 21.7 | 0.04*** | 0.01 | 0.40*** | 0.10 | 0.25** | 0.10 |
| **Calendar years** |  |  |  |  |  |  |  |  |
| 2000 to 2012 | 0 |  | 0 |  | 0 |  | 0 |  |
| 2013 to 2020 | -16.3 | 30.2 | 0.02 | 0.02 | 0.41** | 0.14 | -0.85*** | 0.15 |
| **R<sup>2</sup>** | 21.0 % |  | 4.5 % |  | 13.1% |  | 19.81% |  |

\* p<0.05, \*\*p<0.01, \*\*\*p<0.00

We also plotted predicted curves for neonatal measurements across gestational age 32-42 weeks from the multiple linear regression models. Figure 3 clearly shows that neonates born to mothers with type 1 diabetes had the largest measurements, followed by neonates born to type 2 diabetes, and neonates born to GDM mothers. Neonates of NGT mothers were the smallest.

**Figure 3.**
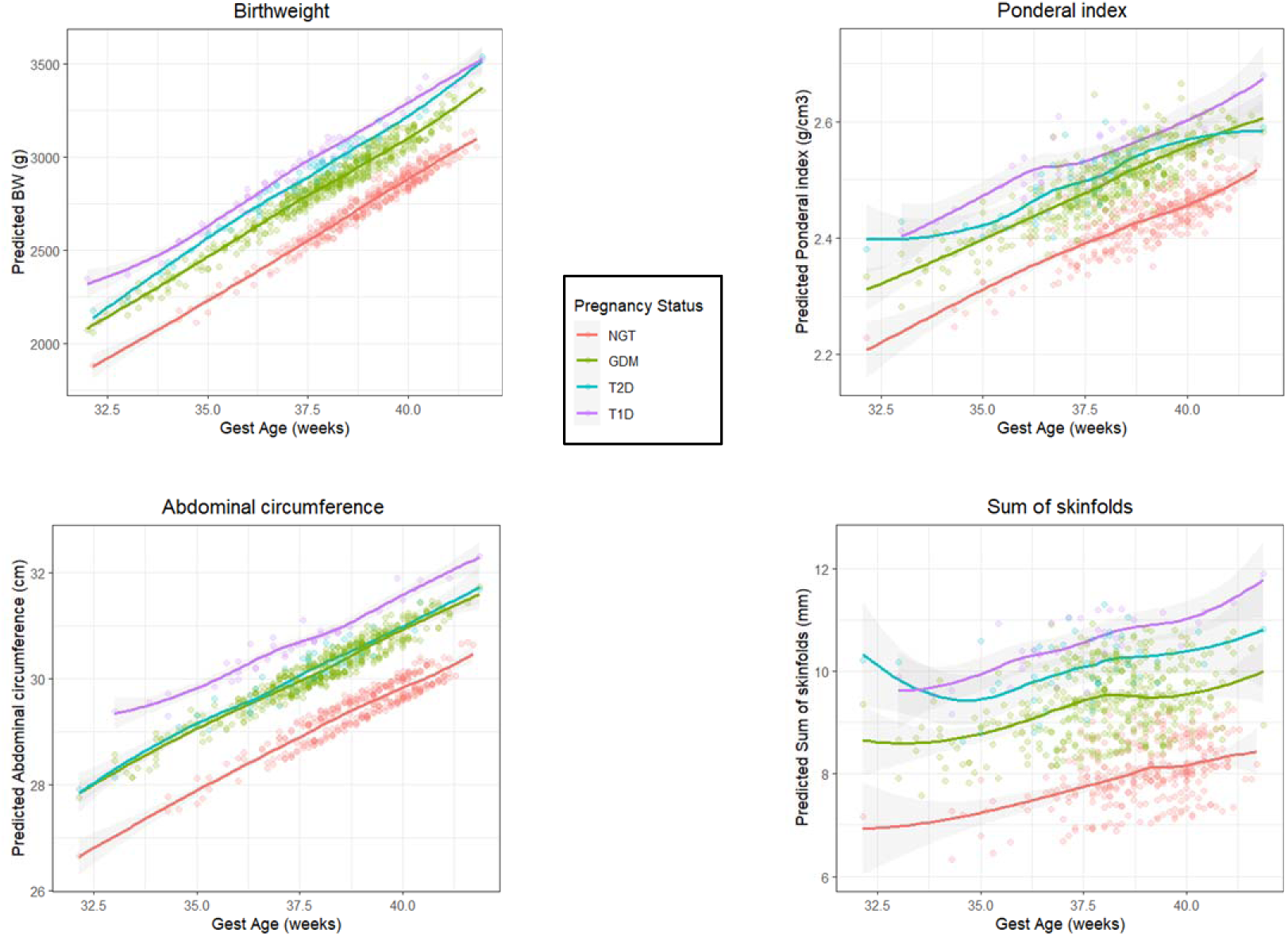
Prediction curves for adiposity measurements in neonates born to mothers with diabetes (type 1, type 2 and GDM) or NGT between gestational age 32 and 42 weeks. Prediction equation included following co-variates: maternal age, neonatal sex, gestational age at birth, parity, and birth year.

### Additional analyses

Step-wise multiple linear regression showed that the effect of maternal type of diabetes was greater than that of pre-pregnancy BMI and GWG on all neonatal adiposity measures. For subcutaneous adiposity (R^2^ change: maternal type of diabetes 11.8 %, maternal pre-pregnancy BMI 0.9%). Comparable results were obtained for ponderal index and abdominal circumference (Supplemental table S5).

## DISCUSSION

In this analysis of clinical data from more than a thousand pregnancies treated in one hospital in India, we confirm that neonates born in pregnancies with diabetes (type 1, type 2 and GDM) are more obese-adipose than those born in normoglycemic pregnancies. Our seminal observation is that the neonates of the least obese-adipose but the most hyperglycemic mothers with type 1 diabetes were the most obese-adipose. Neonates born to mothers with type 2 diabetes and GDM were less so. Our findings in this population are strongly supportive of Pedersen’s ‘hyperglycemia-hyperinsulinemia’ and Freinkel’s ‘fuel-mediated teratogenesis’ models [3,4]. This suggests that maternal hyperglycemia is a much stronger driver of neonatal adiposity than maternal size. This clear finding owes to a distinct dissociation of maternal size and hyperglycemia in our patients. On multivariable regression analysis, we found that type 1 diabetes had the largest effect on neonatal weight, abdominal circumference, and skinfolds (Table 2, Figure 3). Neonates born to mothers with type 1 diabetes were 370 g heavier, those born to mothers with type 2 diabetes 265g heavier, and those of GDM 200g heavier, compared to those born to NGT mothers adjusting for differences in maternal age and parity, neonatal sex and gestational age at delivery. Similar results were seen for neonatal ponderal index, abdominal circumference, and skinfold thickness. Maternal pre-pregnancy BMI and GWG (data available in smaller numbers) had only a modest influence (Supplementary Table S4). When only full-term babies were considered, the results were similar (Supplementary Table S5).

Previous reports are based predominantly on observations in offspring of mothers with diabetes in western populations in whom obesity-adiposity is very prevalent [10], and therefore the individual contributions of these two related but distinct exposures are difficult to separate. A review of the literature shows considerable heterogeneity in the reporting of exposures and outcomes in such studies. Despite the heterogeneity, it is possible to construct a broad picture of these associations and define gaps in the current knowledge. In the EFSOCH (Exeter Family Study of Childhood Health) study of non-diabetic Caucasian mothers (28 wk FPG 4.3 mmol/L, pre-pregnancy BMI 27.8 kg/m^2^), both maternal glycemia and size (adjusted for each other) were positively associated with neonatal weight, length, and BMI [9]; the effect of FPG was much higher compared to BMI (Beta 0.510, 0.038 respectively, for birth weight as an outcome). In the GUSTO study (Growing Up in Singapore Towards healthy Outcomes) of a South Asian population (28 wk FPG 4.4 mmol/L, pre-pregnancy BMI 23.3 kg/m^2^), maternal glycemia (adjusted for BMI) showed a positive association with neonatal abdominal adiposity (measured by MRI) [18]. In the Pune Maternal Nutrition Study (PMNS), a cohort of undernourished non-diabetic rural Indian mothers (28 wk FPG 4.0 mmol/L, pre-pregnancy BMI 18.0 kg/m^2^), maternal FPG and circulating lipids, adjusted for maternal BMI, were associated with neonatal weight and abdominal circumference [19]. Thus, in non-diabetic pregnancies, maternal glycemia, lipids and size influence neonatal obesity-adiposity, usually with the strongest effect of glycemia.

In the Hyperglycemia and Adverse Pregnancy Outcome (HAPO) Study, maternal BMI (adjusting for glycemia) and glycaemia (adjusting for BMI) were both associated with neonatal weight and adiposity (skinfolds and estimated percent body fat) [10, 20, 21], also when women were classified NGT and GDM by IADPSG criteria retrospectively [22]. It is noteworthy that mothers in the HAPO study had a mean BMI of 27.7 kg/m^2^, much higher than in our study. Our earlier study showed that neonates of mothers with GDM had greater length, abdominal circumference, and skinfolds than those born in non-diabetic pregnancies, adjusting for maternal BMI [23]. Similar findings were reported in a study from Mysore, India [24]. Thus, studies in hyperglycaemic pregnancies, maternal size and glycemia showed an independent and additive influence on neonatal obesity-adiposity. However, these studies did not report relative contributions of maternal glycemia and size on neonatal outcomes. An American study showed that neonatal adiposity was largely predicted by maternal glycemia [25].

Type of maternal diabetes in our study was a surrogate for the degree of hyperglycemia and maternal size. Mothers with type 1 diabetes were the most hyperglycemic, those with type 2 diabetes had intermediate and GDM had the least hyperglycemia. Interestingly, mothers with type 1 diabetes were the thinnest and least adipose (BMI, waist circumference, and skinfolds), while both type 2 diabetes and GDM were overweight-adipose. In addition, we constructed predictive models for neonatal obesity-adiposity. Figure 3 demonstrates that neonates born to mothers with type 1 diabetes were the largest for all the adiposity measurements across the range of gestational age commonly encountered in clinical practice (32-42 weeks). We provide clean evidence that maternal hyperglycemia rather than her size had a greater influence on neonatal adiposity.

Most studies have not reported sub-types of maternal diabetes in such associations. Studies in the UK reported that irrespective of the type, maternal diabetes in pregnancy was associated with larger offspring birth size [26–28]. Another study in Canada reported that maternal diabetes (pre-gestational and gestational combined) was associated with greater risk for LGA in the offspring [29]. In these studies, mothers with pregestational diabetes had a pregnancy BMI of 25 kg/m^2^ and those with GDM 27 kg/m^2^.

Studies that report on type of maternal diabetes (type 1 diabetes, type 2 diabetes, and GDM) as distinct exposures show results that are similar to ours, however, all these studies report only on neonatal weight. In the Kaiser Permanente (Southern California) study, The Environmental Determinants of Diabetes in the Young (TEDDY) study, and a Norwegian study neonates born in pregnancies with type 1 diabetes were the heaviest, followed by those born in type 2 diabetes and GDM pregnancies [30–32]. Interestingly, mothers in these studies had considerably higher BMIs than in our study, thus highlighting the primacy of glycemic influence even at higher maternal BMI. We have additionally reported on ponderal index, abdominal circumference, and skinfolds, expanding the observations to neonatal body composition than only weight.

Fetal growth and body composition is influenced by genetic factors (for example FTO gene), and maternal nutrition, metabolism, and hormones which exert epigenetic influences. Our results suggest that maternal hyperglycemia is a major metabolic driver of neonatal adiposity. This is known to act by stimulating fetal hyperinsulinemia which is a major growth hormone in fetal life [3]. Maternal and fetal insulin-like growth factor (IGF-1) levels are increased in pregnancies with diabetes, especially so in type 1 diabetes. IGF-1 acts on placental IGF-1 receptor and promotes fetal overnutrition and overgrowth [33,34]. Figure 4 summarizes relative influence of maternal size (BMI) and glycemia (type of maternal diabetes) on neonatal adiposity in our study, highlighting the role of hyperinsulinemia and IGF system in these associations. Freinkel highlighted an additional role for maternal lipids and amino acids in fetal adiposity [4]. We do not have these measurements in this study but have reported associations of maternal lipids with offspring birth weight and adiposity in the PMNS [19] and InDiaGDM studies [35]. Additionally, vascular dysfunction in type 1 diabetes may cause poor oxygen supply (hypoxia) to the fetoplacental unit, promoting increased angiogenesis and improved placentation [36]. A stereologic study of placentae showed an increased terminal villous volume and surface area from pregnancies in both type 1 and type 2 diabetes, while there was an additional increase in capillary volume, surface, and length in placentae from pregnancies with type 1 diabetes, promoting fetal overnutrition [37].

**Figure 4:**
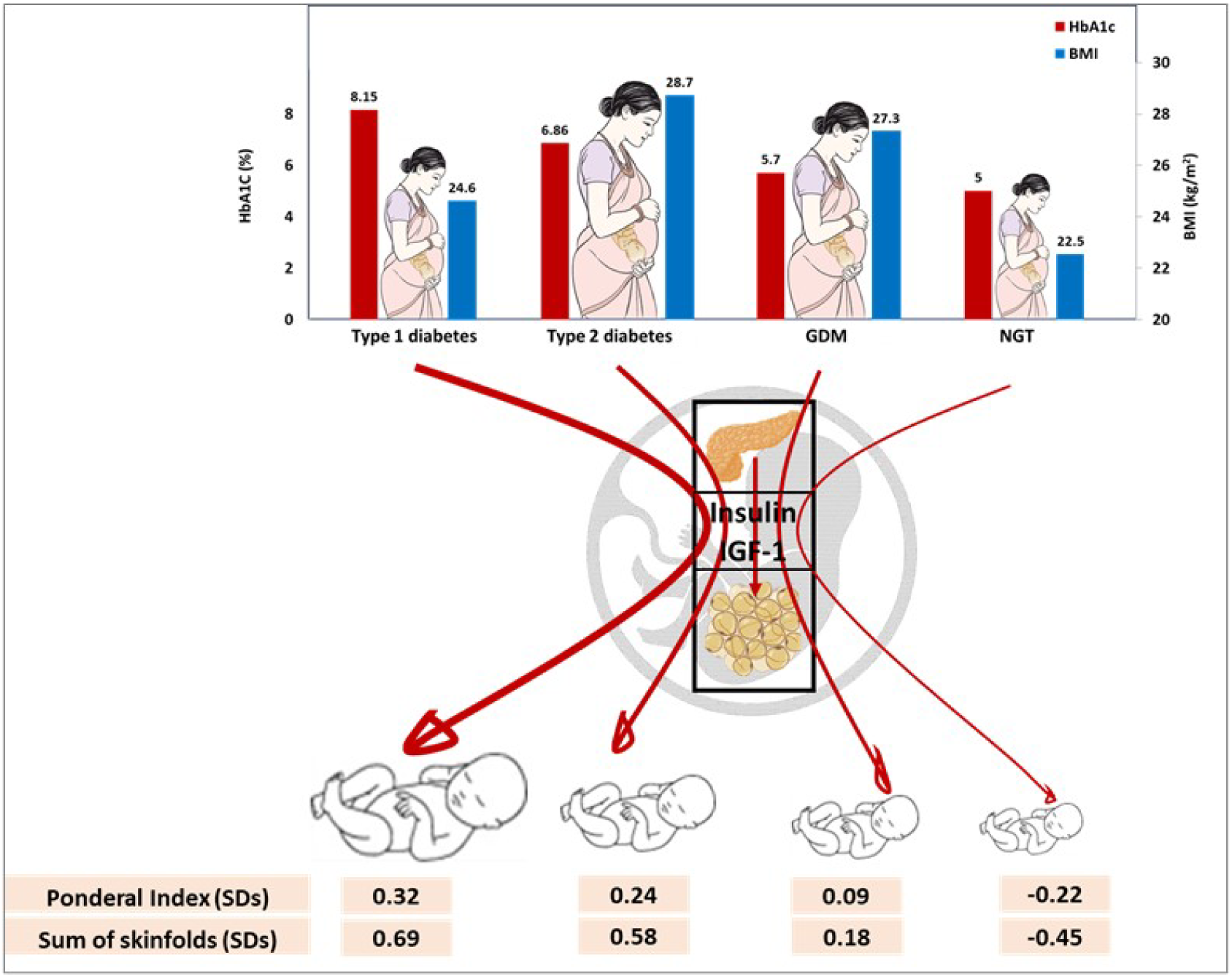
Relative influence of maternal size (BMI) and glycemia (type of maternal diabetes) on neonatal adiposity. Figure shows comparative BMI and glycemia of mothers in our study. Mothers with type 1 diabetes were the most hyperglycemic but the thinnest, mothers with type 2 diabetes and GDM were less hyperglycemic but more overweight-obese. In contrast, neonates of mothers with type 1 diabetes were the most adipose. Mothers with NGT and their neonates were the smallest. Maternal hyperglycemia acts by promoting fetal hyperinsulinemia, which promotes excessive growth of insulin sensitive tissues, especially the adipose tissue. Maternal and fetal IGF-1 and placental IGF-1 receptor also contribute to fetal overgrowth and adiposity.

Finally, studies show that the fetal liver plays a key role in growth regulation and fat deposition. In pregnancies with pre-gestational diabetes, an increase in umbilical flow to the liver in combination with hyperglycemia augments fetal growth [38]. Interestingly, even the small and thin Indian babies are relatively more adipose than English babies [39]. This may result from maternal deficiency of proteins and micronutrients coupled with ‘high-normal’ glycemia and dyslipidaemias creating a double burden of malnutrition for the growing fetus [16, 19, 40]. It is tempting to postulate that the excess adiposity of offspring of mothers with type 1 diabetes in our study may be attributed to these factors in addition to the obvious role of severe hyperglycemia-hyperinsulinemia. It will be interesting to investigate the effects of these metabolic-nutritional factors on the specific steps in adipocyte development from mesenchymal stem cells (lineage commitment, differentiation, and proliferation) [41]. Recently, we have reported a possible role for maternal adipocyte-derived exosomal miRNAs in influencing fetal adiposity [42]. This relationship was partly driven by maternal diabetes. Further research is indicated in this exciting area.

### Strengths and limitations

There are strengths and limitations of this study. We report findings from clinical data on substantial numbers of common subtypes of diabetes in pregnancy, treated in one clinic over 20 years. The GDM and control (NGT) mothers in the study were defined by a 75 g fasting OGTT. Maternal BMI and HbA1c measurements were available in representative numbers, thus helping to ascribe size and glycemic phenotype to the groups (type 1 diabetes, type 2 diabetes, GDM). This allowed us to appreciate disassociation between maternal BMI and glycemia. Measurement of varied adiposity measurements in the offspring allowed definition of body composition in addition to size.

There are some limitations to our study. Given the long duration of the study, and a busy clinical practice, there was sizable missing data. However, our findings of group differences in mothers by type of diabetes are unlikely to be significantly affected. Data on serial blood glucose concentrations, HbA1c in different trimesters, infant breast feeding and paternal size and glycemia was not available. It may be argued that the characteristics of mothers in our study, may not be generalizable to other populations. However, these very unique characteristics provided us with an opportunity to investigate the independent effect of maternal size and hyperglycemia on neonatal obesity-adiposity in a ‘clean’ mechanistic model. Diagnostic criteria for GDM and standards of clinical practice changed during the study period which may introduce some heterogeneity. We have taken account of the variation by including the period of data collection in our linear regression models, and the associations persist after adjustment.

In summary, we confirm increased neonatal adiposity in Indian offspring born in pregnancies with diabetes. Our findings clearly show that maternal hyperglycemia, not the BMI is the major driver of neonatal obesity-adiposity during intrauterine life. This suggests that strict glycaemic control before and during pregnancy will help reduce neonatal adiposity and possibly childhood adiposity in these children.

## Data Availability

Prof. C.S.Yajnik is the guarantor of this work and as such had full access to all the data in the study and takes responsibility for the integrity of the data and the accuracy of the data analysis. Data can be requested from Prof. C.S.Yajnik by applying with a 200 word plan of analysis, data sharing is subject to KEMHRC Ethics Committee approval and Govt. of India Health Ministry Screening Committee permission.

## ABBREVIATIONS

EFSOCH: Exeter Family Study of Childhood Health study
EQAS: external quality control
FPG: Fasting Plasma Glucose
GDM: gestational diabetes mellitus
GUSTO: Growing Up in Singapore Towards healthy Outcomes study
HAPO: Hyperglycemia and Adverse Pregnancy Outcome Study
IADPSG: International Association of Diabetes and Pregnancy Study Groups
IAEA-B12: International Atomic Energy Agency study
IGF: Insulin-like Growth Factor (IGF)
KEMHRC: King Edward Memorial Hospital Research Centre
LGA: large for gestational age
NGT: normal glucose tolerance
OADs: oral anti-diabetic drugs
PMNS: Pune Maternal Nutrition Study
SGA: small for gestational age
TEDDY: The Environmental Determinants of Diabetes in the Young study

## ACKNOWLEDGEMENTS

We are grateful to Prof. Patrick Catalano and Prof. Caroline Fall for informative discussions. We also acknowledge Dr K.M. Shelgikar, Dr. Shailaja Kale, Pallavi Yajnik, Rasika Ladkat, Sonali Rege, Neelam Memane, Vidya Gokhale, and Swati Alekar for their contributions to the study.

## DATA AVAILABILIY

Data can be made available on a reasonable request to the corresponding author, after obtaining appropriate permissions.

## AUTHORS’ RELATIONSHIPS AND ACTIVITIES

CSY was a visiting professor at the Danish Diabetes Academy (supported by Novo Nordisk, Denmark) and University of Southern Denmark during the conduct of the study and writing of this article.

None of the authors declare any conflict of interest.

## DECLARATION

Aspects of this work have been presented at the following national and international conferences: RSSDI (2021), DOHaD (2022), Obesity Week (2022), SNEHA (2023), PSG (2023), Diabetes India World Congress (2024).

## AUTHOR CONTRIBUTION STATEMENT

SDJ was involved in conceptualization and study design, analysis, and interpretation of data, drafting and reviewing the article. SSW and SBP were involved in drafting and reviewing the article. DSB was involved in acquisition of data and reviewing the manuscript, MKD, HGL, SRO, MK, SND, RPK, SGW, DAR, were involved in acquisition of data. RHW and SB were involved in data analysis, CSY was involved in conceptualization and design, interpretation of data, drafting and reviewing the article critically for important intellectual content. All authors approved the final version to be published.

