## Supplemental Table for "Neonatal Adiposity is Largely Driven by Maternal Hyperglycemia *in utero*: Analysis of Routine Clinical Data from a Diabetes Clinic in India"

Supplemental Table S1: Methodology adopted for anthropometric measurements

|  | **Instrument and make** | **Least count** | **Neonate** | **Mother** | **Site of measurement** |
| --- | --- | --- | --- | --- | --- |
| Birth weight (g) | Electronic weighing scale  (ATCO Healthcare Ltd, Mumbai, India) | 50 g | ✓ | X | - |
| Crown heel length | Pedobaby  (ETS J.M.B., Brussels, Belgium) | 0.1 cm | ✓ | X | - |
| Ponderal Index (g/cm^3^)  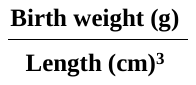 | See above | - | ✓ | X | - |
| Weight (Kg) | Electronic weighing scales  (ATCO Healthcare Ltd, Mumbai, India) | 0.01 Kg | X | ✓ | - |
| Height (cm) | Stadiometer (CMS Instruments Ltd, London, UK). | 0.1 cm | X | ✓ | Head (Frankfurt plane) |
| BMI (kg/m^2^)  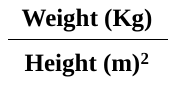 | See above | - | X | ✓ |  |
| Abdominal circumference (cm) | Non- stretchable fiberglass tape (CMS Instruments, London, UK) | 0.1 cm | ✓ | ✓ | Just above the umbilicus |
| Triceps (mm) | Harpenden skinfold calipers (CMS Instruments, London, UK) | 0.1 mm | ✓ | ✓ | The posterior most bulging portion over the tricep at the mid-point between tip of shoulder and tip of olecranon process on the non-dominant arm |
| Biceps (mm) |  |  | ✓ | ✓ | The anterior most bulging portion of upper arm over biceps at the mid-point between tip of shoulder and tip of olecranon process on the non-dominant arm |
| Subscapular (mm) |  |  | ✓ | ✓ | Immediately below the inferior angle of scapula |
| Suprailliac (mm) |  |  | ✓ | ✓ | Upper border of iliac crest is palpated and marked with a pen in the mid-axillary line on the non-dominant side |

Supplementary Figure 1 Participant Flow chart

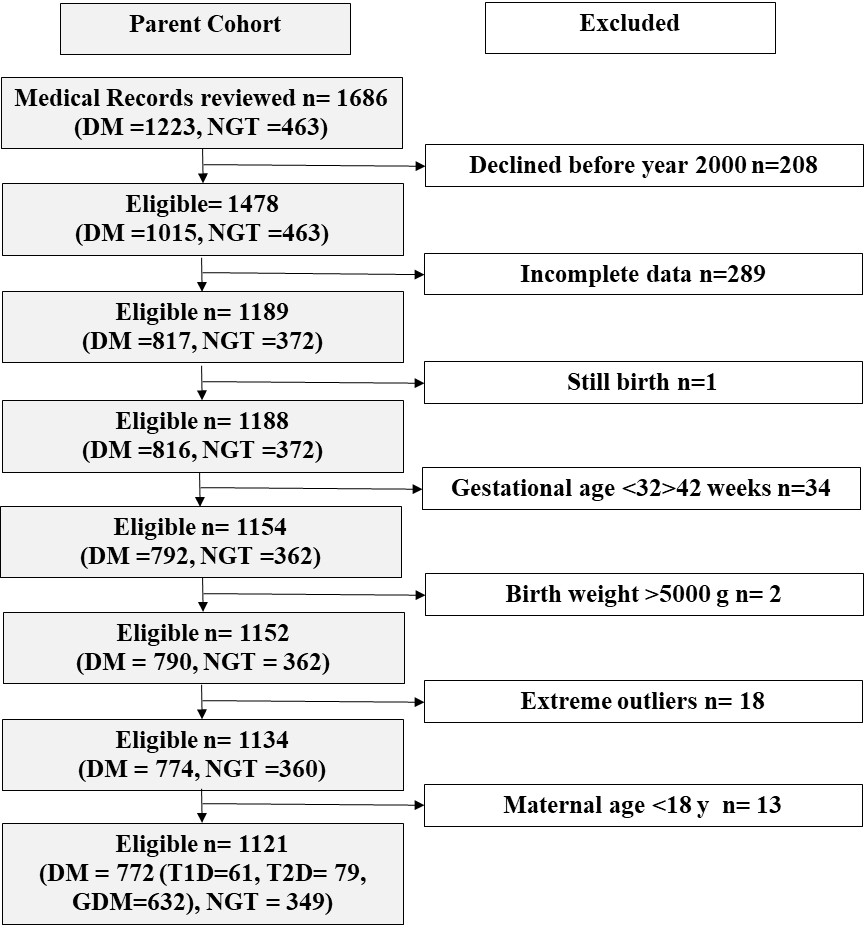

DM; Mothers with diabetes, T1D; mothers with type 1 diabetes; T2D; mothers with type 2 diabetes, NGT; mothers with normal glucose tolerance.

Supplemental Table S2: Maternal and neonatal characters of those excluded and included in the study.

|  | Included n=772 | | Excluded n=243 | |
| --- | --- | --- | --- | --- |
|  | n | Median | n | Median |
| **Maternal characters** |  |  |  |  |
| Age at conception (yrs.) | 626 | 28.6  (25.9, 32.0) | 123 | 28.2  (24.9, 30.9) |
| BMI (kg/m^2^) | 582 | 27.42  (24.6, 30.4) | 96 | 27.52  (24.1, 32.9) |
| Waist circumference (cm) | 393 | 96.0  (89.-0, 102.0) | 79 | 96.3  (91.0, 105.7) |
| Hip circumference (cm) | 391 | 104.00  (98.2, 111.5) | 79 | 104.70  (98.0, 116.0) |
| Biceps skinfold thickness (mm) | 399 | 10.70  (8.1, 14.0) | 80 | 12.07  (8.3, 15.8) |
| Triceps skinfold thickness (mm) | 400 | 23.10  (18.5, 28.3) | 80 | 24.90  (18.3, 29.8) |
| Subscapular skinfold thickness (mm) | 472 | 36.25  (26.8, 44.0) | 94 | 36.55  (28.0, 45.1) |
| Suprailiac skinfold thickness (mm) | 400 | 32.60  (24.9, 41.0) | 80 | 35.55  (27.1, 43.1) |
| Sum of skinfold thickness (mm) | 399 | 102.35  (82.8, 123.8) | 80 | 108.25  (83.5, 127.6) |
| HbA1c (%) | 104 | 6.20  (5.5, 7.6) | 20 | 7.00  (5.6, 9.9) |
| HbA1c (mmol/mol) | 104 | 44.0  (37.0, 60.0) | 20 | 53.0  (38.0, 85.0) |
| **Neonatal characters** |  |  |  |  |
| Gestational age (weeks) | 772 | 38.0  (36.9, 39.0) | 127 | 37.6***  (34.1, 38.7) |
| Gender (Male) (n %) | 772 | 425 (55.1%) | 193 | 105 (43.2%) |
| Birth weight (kg) | 772 | 2.9  (2.5, 3.2) | 139 | 2.9  (2.3, 3.2) |
| Length (cm) | 589 | 48.9  (47.1, 50.0) | 77 | 48.3  (47.0, 50.0) |
| Head circumference (cm) | 595 | 33.4  (32.3, 34.3) | 78 | 33.4  (32.5, 34.3) |
| Abdominal circumference (cm) | 594 | 30.1  (28.6, 31.6) | 77 | 30.4  (28.8, 32.3) |
| Triceps skinfold thickness (mm) | 591 | 4.3  (3.6, 5.4) | 77 | 4.6  (4.4, 6.2) |
| sub-scapular skinfold thickness (mm) | 588 | 4.9  (4.0, 6.0) | 77 | 5.3  (4.4, 6.2) |
| Sum of skinfold thickness (mm) | 588 | 9.3  (7.8, 11.1) | 77 | 10.1  (8.4, 11.5) |

Data shown as median (25^th^, 75^th^ percentiles). * p<0.05, **p<0.01, ***p<0.001

Supplemental Table S3: Characters of mothers and neonates where maternal gestational weight gain data was available or not available.

|  | GWG not available (n=495) | | GWG available  (n=277) | |
| --- | --- | --- | --- | --- |
|  | N | Median | N | Median |
| Age (yrs.) | 363 | 29.0  (26.0, 32.7) | 263 | 28.0*  (25.3, 31.5) |
| Weight at delivery (kg) | 333 | 66.0  (57.8, 72.8) | 277 | 64.9  (57.9, 73.8) |
| Height (cm) | 371 | 155.0  (151.3, 158.5) | 274 | 155.1  (151.3, 158.8) |
| BMI at delivery (kg/m^2^) | 324 | 27.3  (24.5, 30.1) | 274 | 27.0  (24.2, 30.0) |
| Sum of skinfold thickness (mm) | 254 | 98.5  (82.1, 118.9) | 145 | 105.5*  (84.3, 126.2) |
| HbA1c (%) | 72 | 6.1  (5.3, 7.3) | 31 | 6.8*  (5.9, 8.0) |
| HbA1c (mmol/mol) | 72 | 43.0  (34.0, 56.0) | 31 | 51.0*  (41.0, 64.0) |
| Gestational age (weeks) | 495 | 37.9  (36.6, 38.9) | 277 | 38.1**  (37.1, 39.1) |
| Birth weight (kg) | 495 | 2.8  (2.5, 3.2) | 277 | 3.0  (2.6, 3.2) |
| Length (cm) | 331 | 48.9  (47.1, 50.2) | 258 | 49.0  (47.3, 50.0) |
| Abdominal circumference (cm) | 334 | 30.0  (28.5, 31.6) | 260 | 30.4  (29.0, 31.9) |
| Sum of skinfold thickness (mm) | 330 | 9.3  (8.0, 11.6) | 258 | 9.2  (7.6, 10.8) |

Data shown as median (25^th^, 75^th^ percentiles). * p<0.05, **p<0.01, ***p<0.001

Supplemental Table S4: Multiple linear regression results to study association between maternal type of diabetes and neonatal adiposity - analysis on sub set of full-term neonates.

|  | **Birth weight**  **(g) (n=733)** | | **Ponderal Index (g/cm^3^) (n=531)** | | **Abdominal circumference (cm) (n=545)** | | **Sum of skinfolds (mm) (n=533)** | |
| --- | --- | --- | --- | --- | --- | --- | --- | --- |
|  | **Estimate** | **SE** | **Estimate** | **SE** | **Estimate** | **SE** | **Estimate** | **SE** |
| **Type of diabetes** | | | | | | | | |
| Nil (NGT) | 0 | | 0 | | 0 | | 0 | |
| Type 1 diabetes | 281.8 | 88.4 | 0.09 | 0.07 | 1.6*** | 0.4 | 1.8*** | 0.4 |
| Type 2 diabetes | 232.7*** | 71.5 | 0.07 | 0.05 | 0.8** | 0.4 | 1.5*** | 0.4 |
| GDM | 192.3*** | 35.2 | 0.10*** | 0.02 | 1.1*** | 0.2 | 1.4*** | 0.2 |
| **Maternal age (yrs)** | 2.2 | 3.8 | 0.0 | 0.0 | 0.0 | 0.0 | 0.0 | 0.0 |
| **Gestational age (wk)** | 69.5*** | 13.7 | 0.02 | 0.01 | 0.2*** | 0.07 | -0.01 | 0.0 |
| **Sex of neonate** | | | | | | | | |
| Male | 0 | | 0 | | 0 | | 0 | |
| Female | -60.3 | 29.1 | 0.03 | 0.02 | -0.1 | 0.1 | 0.6*** | 0.1 |
| **Parity** | 49.2 | 22.6 | 0.03 | 0.02 | 0.30 | 0.1 | 0.1 | 0.11 |
| **Calander years** | | | | | | | | |
| 2000 to 2012 | 0 | | 0 | | 0 | | 0 | |
| 2013 to 2020 | -6.1 | 30.6 | 0.03 | 0.02 | 0.4 | 0.1 | -0.9*** | 0.1 |
| **R^2^** | 8.3% | | 2.6% | | 8.5% | | 19.8% | |

* p<0.05, **p<0.01, ***p<0.001

Supplemental Table S5: Multiple stepwise regression analysis for factors affecting neonatal adiposity

| **Neonatal outcome** | **Factor** | **r^2^** | **∆ r^2^** | **p value** |
| --- | --- | --- | --- | --- |
| **Birth Weight** | Gestational age | 0.096 | 0.096 | 0.000 |
|  | Maternal diabetes Type | 0.138 | 0.042 | 0.000 |
|  | Maternal pre-pregnancy BMI | 0.148 | 0.010 | 0.020 |
|  | Gestational weight gain | 0.157 | 0.008 | 0.032 |
| **Ponderal index** | Maternal pre-pregnancy BMI | 0.015 | 0.015 | 0.010 |
|  | Gestational weight gain | 0.028 | 0.013 | 0.015 |
| **Abdominal circumference** | Gestational age | 0.032 | 0.032 | 0.000 |
|  | Maternal diabetes Type | 0.084 | 0.052 | 0.000 |
|  | Maternal pre-pregnancy BMI | 0.101 | 0.017 | 0.004 |
|  | Gestational weight gain | 0.112 | 0.011 | 0.019 |
|  | Parity | 0.120 | 0.008 | 0.039 |
| **Sum of skinfolds** | Maternal diabetes Type | 0.118 | 0.118 | 0.000 |
|  | Neonatal sex | 0.142 | 0.024 | 0.000 |
|  | Maternal pre-pregnancy BMI | 0.150 | 0.009 | 0.034 |
